# The effect of Omicron breakthrough infection and extended BNT162b2 booster dosing on neutralization breadth against SARS-CoV-2 variants of concern

**DOI:** 10.1101/2022.08.04.22278160

**Authors:** Carl Graham, Thomas Lechmere, Aisha Rehman, Jeffrey Seow, Ashwini Kurshan, Isabella Huettner, Thomas J.A. Maguire, Jerry Tam, Daniel Cox, Christopher Ward, Mariusz Racz, Anele Waters, Christine Mant, Michael H. Malim, Julie Fox, Katie J. Doores

**Affiliations:** Department of Infectious Diseases, School of Immunology & Microbial Sciences, King’s College London, London, UK; Harrison Wing, Guys and St Thomas’ NHS Trust, London, UK; Infectious Diseases Biobank, Department of Infectious Diseases, School of Immunology and Microbial Sciences, King’s College London, London, UK

## Abstract

COVID-19 vaccines are playing a vital role in controlling the COVID-19 pandemic. As SARS-CoV-2 variants encoding mutations in the surface glycoprotein, Spike, continue to emerge, there is increased need to identify immunogens and vaccination regimens that provide the broadest and most durable immune responses. We compared the magnitude and breadth of the neutralizing antibody response, as well as levels of Spike-reactive memory B cells, in individuals receiving a second dose of BNT126b2 at a short (3-4 week) or extended interval (8-12 weeks) and following a third vaccination approximately 6-8 months later. We show that whilst an extended interval between the first two vaccinations can greatly increase the breadth of the immune response and generate a higher proportion of Spike reactive memory B cells, a third vaccination leads to similar levels between the two groups. Furthermore, we show that the third vaccine dose enhances neutralization activity against omicron lineage members BA.1, BA.2 and BA.4/BA.5 and this is further increased following breakthrough infection during the UK omicron wave. These findings are relevant for vaccination strategies in populations where COVID-19 vaccine coverage remains low.

## Introduction

Development of vaccines against SARS-CoV-2 has been a vital step in controlling the global COVID-19 pandemic. Most approved vaccines use the SARS-CoV-2 Spike antigen to elicit a neutralizing antibody response as well as generating cell-mediated immunity. The Spike glycoprotein interacts with the angiotensin-converting enzyme 2 (ACE2) on host cells and facilitates viral entry. One of the greatest challenges faced by current vaccines has been the emergence of SARS-CoV-2 viral variants of concern (VOCs) that encode mutations in the Spike protein, including alpha (B.1.1.7), beta (B.1.351), delta (B.1.617.2) and omicron (sub-lineage members BA.1, BA.2, BA.4 and BA.5). Numerous studies have shown that these Spike mutations can lead to partial escape in convalescent and vaccinee sera, with the greatest reduction in neutralization being observed for omicron sub-lineages [1-5]. Therefore, immunogens and immunization regimes that elicit durable neutralizing antibody (nAb) responses with broad activity against both known and newly emerging variants is highly desirable.

In December 2020, the Pfizer/BioNTech BNT162b2 mRNA COVID-19 vaccine was given approval for use in the UK. The approved regimen was two doses administered at a 3– 4-week interval, but in January 2021 a change to UK policy meant that the BNT162b2 booster vaccination was administered with an extended interval of 8–12 weeks. The rationale behind this change was to provide as much of the UK population with some level of immunity in the face of a large wave of SARS-CoV-2 alpha variant infections [6]. Initially, most COVID-19 vaccines required two vaccinations to provide efficacy against the Wuhan (wild-type, WT) alpha and delta VOCs. However, due to waning of vaccine-elicited nAb levels [7] and with the emergence of omicron/BA.1, which encodes >30 mutations in Spike, a third vaccination (either BNT162b2 or mRNA-1273) became recommended. A third vaccine has been shown to increase neutralization titres against omicron/BA.1 [3-5, 8] and to restore vaccine efficacy against omicron/BA.1 [9-11] however, breakthrough infections (BTI) in vaccinated individuals have been observed at increased frequencies compared to previous dominant variants [9-11].

We sought to determine how the interval between the 1^st^ and 2^nd^ BNT162b2 vaccinations impacted on neutralization breadth and potency against current and newly emerging VOCs in the short-term (post 2-doses) and in the long-term (post 3-doses), and how subsequent BTI during the UK omicron wave (January to February 2022) affected neutralization of newer omicron sub-lineage members. We show that broader plasma neutralizing activity is observed when the 2^nd^ dose is given with extended 8–12 week interval (extended group) compared to those receiving the 2^nd^ dose with a 3-4 week interval (short group). However, the advantage of the extended booster to neutralization is not displayed following a 3^rd^ vaccination where robust neutralization of omicron lineages members (BA.1, BA.2, BA.4 and BA.5) is observed in both groups. Finally, we show that omicron BTI further boosts the neutralization activity against omicron sub-lineages. Overall, this research provides insights into optimizing vaccine regimens to provide the greatest neutralization breadth.

## Results

### Vaccine cohort description

Plasma and PBMCs were collected from individuals receiving the BNT162b2 vaccine either with a short booster interval (3-4 weeks, n = 19, short-group) or an extended booster interval (8-12 weeks, n = 28, extended-group) (**Figure 1A**). For the extended-group blood was collected prior to vaccination (visit 1), 3 weeks post 1^st^ dose (visit 2) and post 2^nd^ dose (visit 3). For the short-group, blood was only collected post 2^nd^ dose (visit 3). Blood was also collected from both groups 6 months post 2^nd^ vaccine dose (visit 4) and further samples was taken 3-4 weeks and 6-months post 3^rd^ vaccine (visit 5 and visit 6, respectively). For individuals experiencing a BTI, blood was also collected post infection (visit 7).

**Figure 1:**
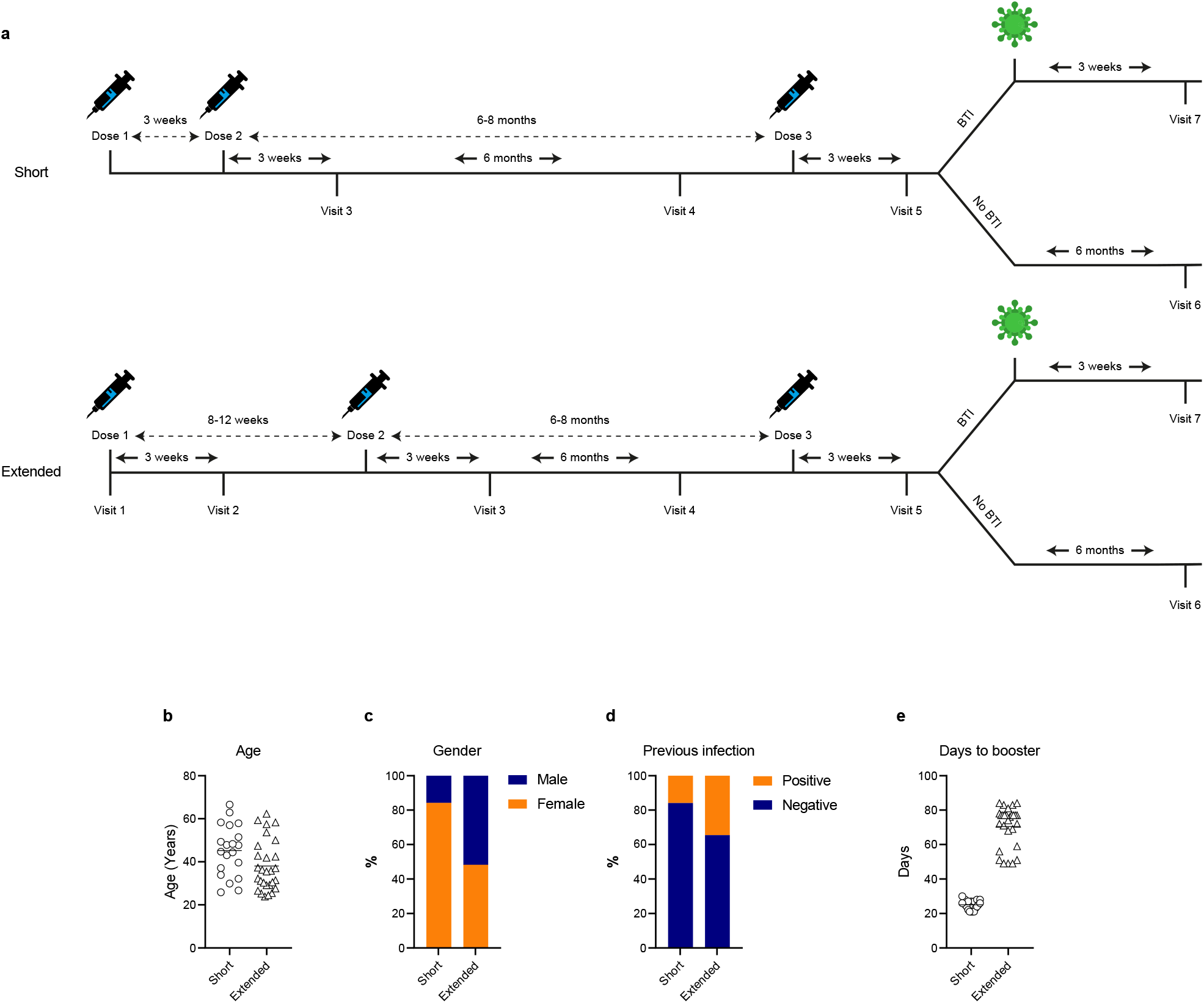
**A)** Vaccination and blood sampling schedule in short and extended vaccine groups. Horizontal line shows the mean age for each group. **B)** Difference in age between short- and extended-groups. **C)** Gender of participants in each group. **D)** Frequency of previous SARS-CoV-2 infection. **E)** Length of time between the 1^st^ and 2^nd^ dose of BNT162b2 vaccine. Horizontal line shows the mean days between vaccine doses. Short- and extended-groups shown in circle and triangles, respectively.

The short-group was 84.2% female, with an average age of 44.1 years (interquartile range, 15.0 years) whereas the extended-group was 53.6% female, with a slightly lower average age of 38.0 years (interquartile range, 14.7 years) (**Figure 1B&C**). Information on previous SARS-CoV-2 infection was recorded at each blood draw and the presence of IgG to Nucleoprotein (N) was measured to determine previous asymptomatic SARS-CoV-2 infections or infections occurring prior to community PCR testing. Previous infection was detected in 3/19 from the short-group and in 10/28from the extended-group (**Figure 1D**). The average interval between vaccine doses was 25.2 days (interquartile range, 3.5 days) for the short group and 71.0 days (interquartile range, 11.3 days) in the extended group (**Figure 1E**).

### Anti-Spike IgG levels are similar in short and extended booster groups

IgG binding to recombinant Wuhan-1 Spike was measured by ELISA and the half-maximal binding (ED_50_) calculated (**Figure 2**). As reported previously [12-16], individuals who had experienced a SARS-CoV-2 infection prior to vaccination had higher Spike IgG levels compared to naïve individuals after the 1^st^ vaccine dose (visit 2) (**Figure 2B**). However, similar levels of anti-Spike IgG were observed in the SARS-CoV-2 naïve and convalescent donors following the 2^nd^ vaccination (visit 3) (**Figures 2A** and **2B**).

**Figure 2:**
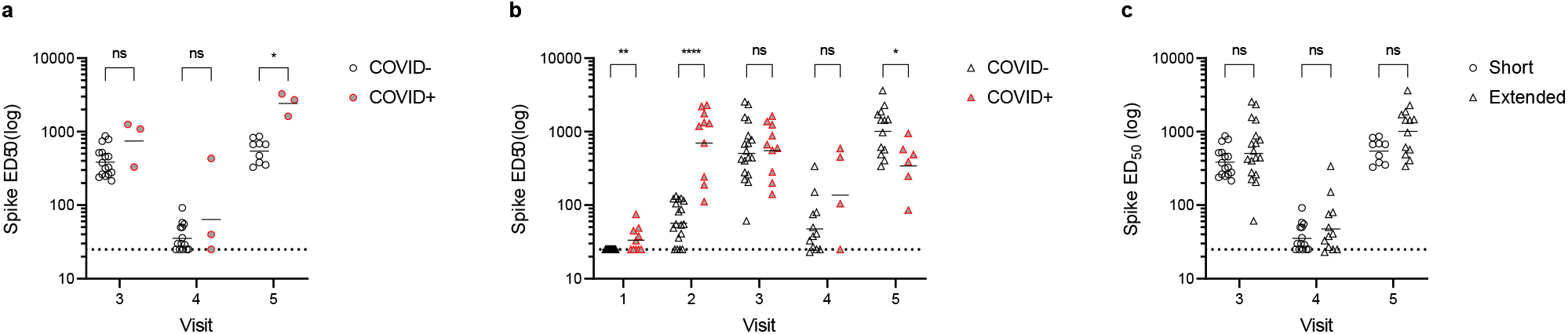
IgG binding titres to SARS-CoV-2 Spike over time. **A)** Comparison of geometric mean ED_50_ values between SARS-CoV-2 naïve individuals (open black circle) and those previously infected before vaccination (grey/red circles) in short-booster group. **B**) Comparison of geometric mean ED_50_ values between SARS-CoV-2 naïve individuals (open black triangle) and those previously infected before vaccination (grey/red triangle) in extended-booster group. **C**) Comparison of geometric mean ED_50_ values between short-and extended-booster groups at visit 3, 4 and 5. Dotted line represents the lowest limit of detection for the assay. D’Agostino and Pearson tests were performed to determine normality. Based on this result, multiple Mann-Whitney tests or unpaired t tests using a two-stage linear stepup procedure of Benjamini, Krieger and Yekutieli were employed to determine significance between groups. ns P > 0.05, * P ≤ 0.05, ** P ≤ 0.01, *** P ≤ 0.001 and **** P ≤ 0.0001.

For SARS-CoV-2 naïve individuals, comparison of ED_50_ between the short and extended groups showed no statistical differences at visit 3, although there was a trend towards higher levels in the extended group (**Figure 2C**). Both groups showed waning of the anti-Spike IgG levels 6 months post 2^nd^ vaccine (visit 4) which were subsequently boosted after participants received a 3^rd^ vaccine dose (visit 5) (**Figures 2A and 2B**).

### Previous SARS-CoV-2 infection leads to higher nAb titres following the first vaccine dose

Next, plasma neutralization breadth and potency were determined using HIV-1 lentiviral particles pseudotyped with either the SARS-CoV-2 Spike of Wuhan-1 (vaccine strain), alpha, delta, beta or omicron/BA.1 VOCs and a HeLa cell line stably expressing ACE2 as the target cell [17]. Analysis of neutralizing responses after a single vaccine dose (visit 2) was only conducted on the extended-group due to sample availability (**Figure 1A**). Consistent with previous studies [12-16], following 1-dose of BNT162b2, geometric mean titres (GMTs) against the matched vaccine strain (Wuhan-1, wildtype, WT) were higher is those who had a SARS-CoV-2 infection prior to vaccination (**Figures 3A-3C**). Furthermore, previously infected individuals showed greater neutralization breadth against VOCs (**Figure 3A-3C**).

**Figure 3:**
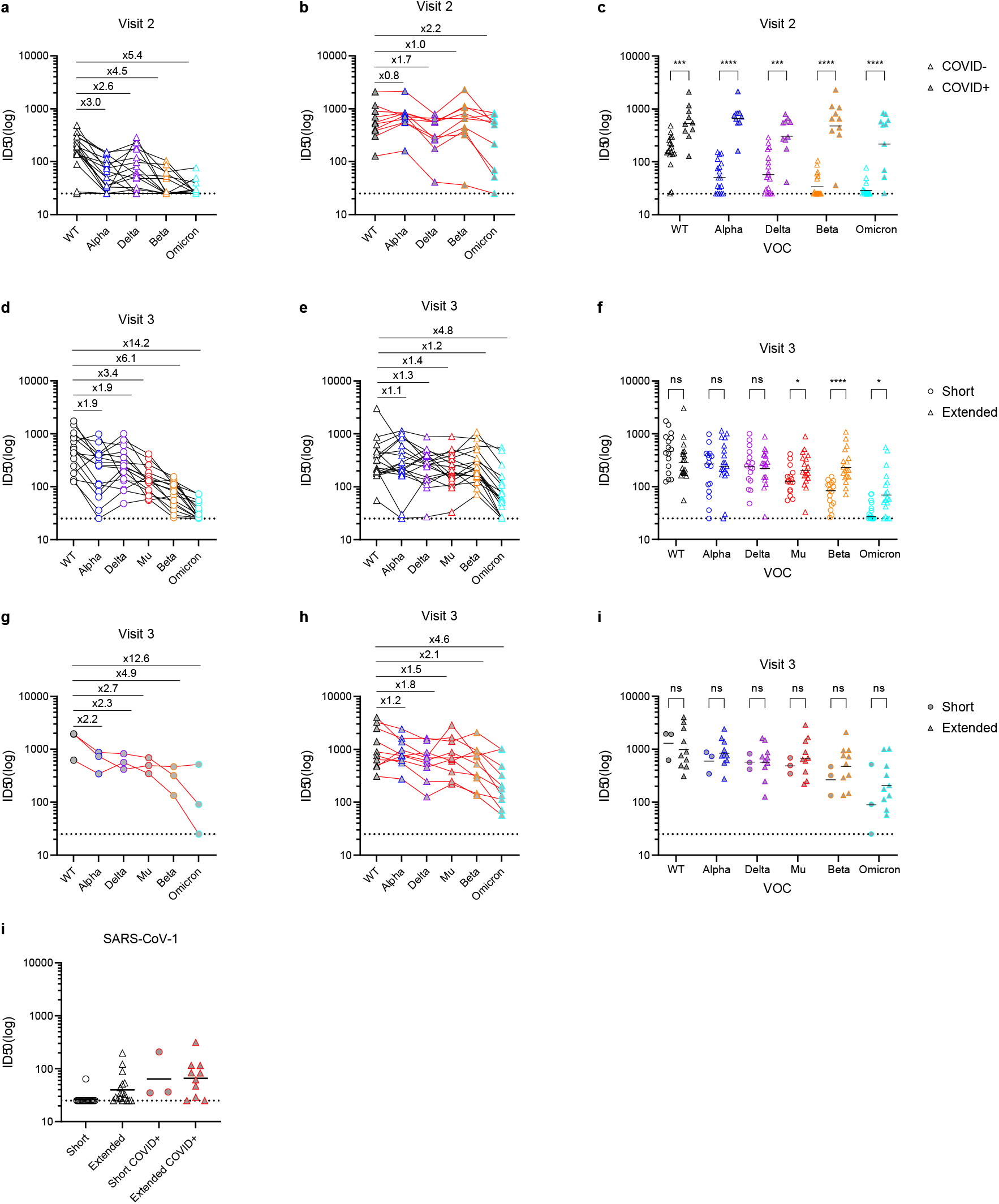
Neutralization breadth and potency following two-doses of BNT162b2. Neutralization was measured against wuhan-1 (WT), alpha, delta, mu, beta and omicron/BA.1. **A)** Neutralization following a single BNT162b2 vaccine dose (visit 2) in naïve individuals (n = 18). **B)** Neutralization following a single BNT162b2 vaccine dose in individuals infected with SARS-CoV-2 prior to vaccination (n = 10). Data from individual donors are linked and fold changes compared to WT shown above. **C)** GMT comparison between SARS-CoV-2 naïve individuals (open triangle) and convalescent donors (grey triangle) following one vaccine dose. The horizontal line shows the GMT. Neutralization data following 2-vaccine doses (visit 3) in SARS-CoV-2 naïve individuals in **D)** the short booster group (n = 16), and **E)** the extended booster group (n = 18). Data from individual donors are linked and fold changes compared to WT shown above. **F)** GMT comparison in SARS-CoV-2 naïve individuals receiving the short booster interval (open circle) and extended booster (open triangle). The horizontal line shows the GMT. Neutralization data following 2-vaccine doses (visit 3) in SARS-CoV-2 convalescent individuals in **G)** the short booster group (n = 3) and **H)** the extended booster group (n = 9). Data from individual donors are linked and fold changes compared to WT shown above. **I)** GMT comparison in SARS-CoV-2 convalescent individuals receiving the short booster interval (grey circle) and extended booster (grey triangle). The horizontal line shows the GMT. **J)** Neutralization of SARS-CoV-1. The horizontal dotted line is the lowest limit of detection. D’Agostino and Pearson tests were performed to determine normality. Based on this result, multiple Mann-Whitney tests using a two-stage linear step-up procedure of Benjamini, Krieger and Yekutieli were employed to determine significance between groups. ns P > 0.05, * P ≤ 0.05, ** P ≤ 0.01, *** P ≤ 0.001 and **** P ≤ 0.0001.

### An extended interval between the 2^nd^ vaccine dose enhances the breadth of the neutralizing antibody response

Following two doses of BNT162b2 in SARS-CoV-2 naïve individuals, GMTs against the matched vaccine strain (Wuhan-1, WT) were similar between the short and extended booster groups (**Figure 3D-3F**). In both the short and extended-groups, higher GMTs were observed for individuals that had a SARS-CoV-2 infection prior to vaccination (**Figure 3G-3I**) [12-16]. In SARS-CoV-2 naïve individuals (**Figures 3D and 3E**), the broadest neutralization activity against VOCs was observed in the extended booster group where a modest 1.1–1.4-fold reduction in neutralization was observed against alpha, delta, mu and beta and a 3.6-fold reduction against omicron/BA.1 (**Figure 3E**). For those in the short-group, a larger reduction in neutralization potency against VOCs was observed, with greater reductions against beta (6.1-fold) and omicron (14.2-fold) (**Figure 3D**). Comparison between GMTs of the short and extended booster groups showed a significantly reduced potency against mu, beta and omicron/BA.1 VOCs in the short booster group (**Figure 3F**). Although overall GMTs against VOCs were higher in previously infected vaccinees (**Figure 3G-3I**), the extended booster group also showed higher GMTs against beta and omicron/BA.1 than the short booster group. However, this difference did not reach significance due to the small sample size (**Figure 3I**).

To determine whether the extended booster generated neutralization breadth beyond SARS-CoV-2 VOCs, neutralization was also measured against SARS-CoV-1 which shares 73% sequence [18] similarity with Spike of SARS-CoV-2. Although neutralization titres were generally low, individuals receiving the extended booster showed low levels of SARS-CoV-1 neutralization (**Figure 3J**) with a GMT that was higher than in the short-group.

### An extended vaccine interval enhances the magnitude of the memory B cell response

To further examine how vaccine interval impacts on the B cell response, we next measured the frequencies of Spike-reactive memory B cells in the short and extended booster groups using flow cytometry (**Figure 4A-B** and **Figure S1A**) [12, 19]. The frequency of Wuhan-1 Spike-reactive memory B cells was measured in pre-vaccination samples (visit 1) for the extended booster group only (due to sample availability) and in both groups following the 2 ^nd^ vaccination (visit 3). Overall, where paired samples were available, the overall frequency of memory B cells did not change over the course of the analysis (**Figure S1B**). Participants who had a SARS-CoV-2 infection prior to vaccination had a distinct population of Spike reactive memory B cells in the pre-vaccine sample (mean 0.20 %, range 0.10 – 0.34%) compared to naïve individuals (mean 0.02 %, range 0.01 – 0.06%) (**Figure 4C**). Following two vaccine doses, the percentage of Spike-reactive memory B cells increased independent of prior SARS-CoV-2 exposure (**Figure 4C**).

**Figure 4.**
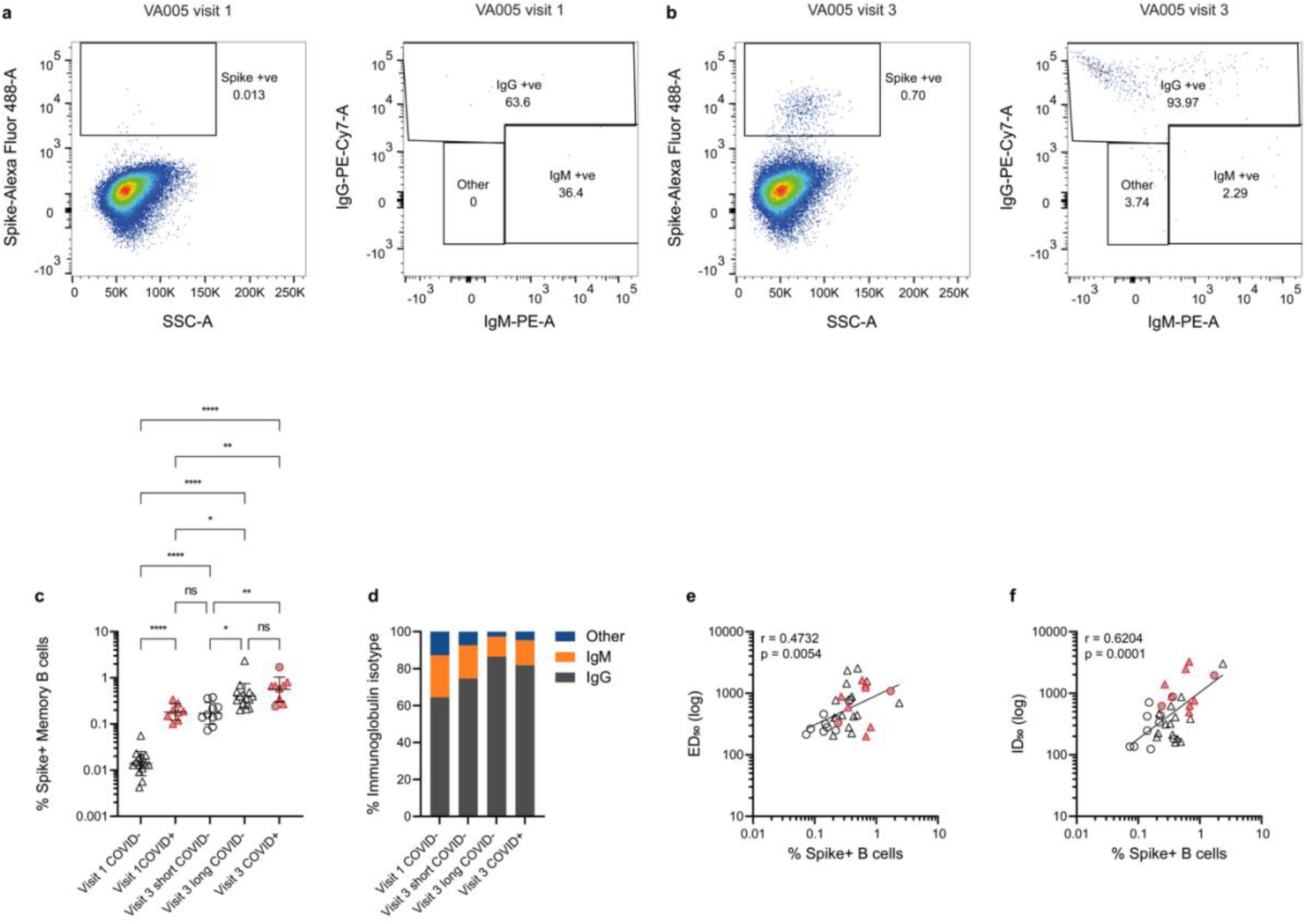
Higher percentage of SARS-CoV-2 Spike-reactive memory B cells are detected in the extended booster group. Percentage of Spike-reactive memory B cells were determined by flow cytometry. **A)** Example pre-bleed (visit 1) PBMC staining for SARS-CoV-2 naïve individual. **B)** Example visit 3 PBMC staining for SARS-CoV-2 naïve individual. Example of full flow analysis shown if **Figure S1A. C)** Percentage Spike-reactive memory B cells at visit 1 and visit 3 were determined by flow cytometry. Previously infected individuals shown in red/grey. Extended and short interval groups shown with triangle and circle symbols, respectively. D’Agostino and Pearson tests were performed to determine normality. Based on this result, differences between groups were assessed using a Brown-Forsythe ANOVA test with Dunnett’s T3 multiple comparisons post hoc. ns P > 0.05, * P ≤ 0.05, ** P ≤ 0.01, *** P ≤ 0.001 and **** P ≤ 0.0001. **D)** Percentage of IgG, IgM or other Spike-reactive memory B cells for each vaccine group. **E)** Correlation between Spike IgG ED_50_ and percentage of Spike-reactive memory B cells. **F)** Correlation between ID_50_ against Wuhan-1 pseudotyped virus and percentage of Spike-reactive memory B cells. (Spearman’s correlation, r; a linear regression was used to calculate the goodness of fit, r^2^).

In SARS-CoV-2 naïve participants, a higher frequency of Spike reactive memory B cells was observed in the extended booster group (mean 0.52 %, range 0.20 – 2.36%) compared to the short booster group (mean 0.19 %, range 0.07 – 0.38%) after the 2^nd^ vaccine dose (**Figure 4C**). When considering the frequency of Spike-reactive B cells in previously infected vaccinated donors, the percentage (mean 0.67 %, range 0.24 – 1.71 %) was similar to SARS-CoV-2 naïve individuals in the extended group.

The isotype of the Spike-reactive memory B cells was considered further (**Figure 4D**). In samples collected from previously infected individuals prior to vaccination, an average 64.5% of the Spike-reactive memory B cells were IgG positive (**Figure 4D**) and the average percentage increased upon vaccination to 81.8% (**Figure 4D**) suggesting an improvement in the quality of the response. In COVID-19 naïve individuals, a higher percentage of IgG+ Spike-reactive memory B cells was seen in the extended-booster group (86.5%) compared to the short-booster group (74.5%). A positive correlation between the percentage of Spike-reactive memory B cells and the ED_50_ and ID_50_ (**Figures 4E and 4F**) was observed after two vaccine doses independent of booster interval or previous SARS-CoV-2 exposure.

Overall, an extended period between the first and second vaccine dose increases the magnitude of the Spike-specific B cell response.

### Neutralizing antibody levels decline at 6 months but are boosted and neutralization broadened by a third vaccine dose

To understand the durability of the nAb response following two vaccine doses, neutralization titres were measured 6 months post second vaccine dose (visit 4). Plasma neutralization activity against all VOCs (WT, alpha, delta, beta and omicron (BA.1)) had declined in both groups (**Figure 5A-D**) as reported in other recent studies [4, 8, 20]. Although the neutralization of the most antigenically distant VOCs (beta and omicron/BA.1) had declined, titres remained higher in the extended booster group compared to the short booster group (**Figure 5A** and **5B**).

**Figure 5.**
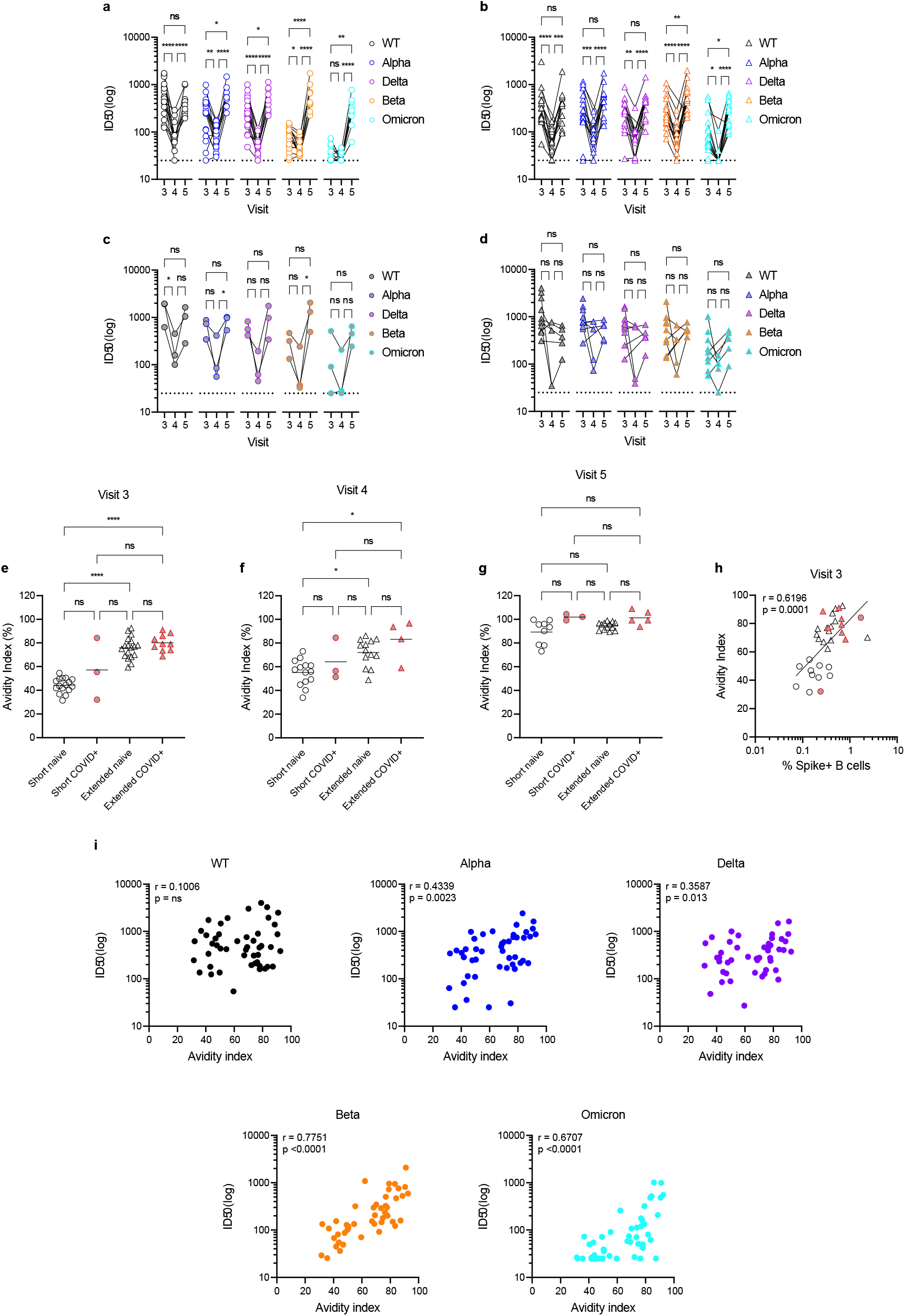
Neutralization potency decreases 6-months post 2^nd^ vaccine but breadth and potency increase after a 3^rd^ vaccine dose. Neutralization ID_50_ against WT, alpha, delta, beta and omicron/BA.1 at visits 3, 4 and 5 in the **A)** short (circle) and **B)** extended (triangle) booster groups for SARS-CoV-2 naïve individuals and **C)** short (grey circle) and **D)** extended (grey triangle) booster groups previously infected individuals. Difference between GMT at visits 3, 4 and 5 were determined using Mann-Whitney unpaired t-test. D’Agostino and Pearson tests were performed to determine normality. Based on this result, differences between groups were assessed using either one-way ANOVA (Tukey’s multiple comparisons post hoc), Kruskal-Wallis test (Dunn’s multiple comparisons post hoc) or Friedman test (Dunn’s multiple comparisons post hoc). ns P > 0.05, * P ≤ 0.05, ** P ≤ 0.01, *** P ≤ 0.001 and **** P ≤ 0.0001. Avidity index for IgG binding to Spike was measured at **E)** visit 3, **F)** visit 4 and **G)** visit 5 and compared across groups. Avidity index was calculated by comparing the area under the curve (AUC) with/without an 8M Urea washing step. D’Agostino and Pearson tests were performed to determine normality. Based on this result, differences between groups were assessed using a Kruskal-Wallis test with Dunn’s multiple comparisons post hoc. **H)** Correlations between avidity index and ID_50_ against VOCs following 2-doses of BNT162b2 vaccine. **I)** Correlation between avidity index and % Spike-reactive memory B cells. (Spearman’s correlation, *r*; a linear regression was used to calculate the goodness of fit, *r*^*2*^).

In September 2021, a vaccine booster programme was initiated in the UK and blood was collected from individuals 3-weeks after receiving a 3^rd^ vaccine dose (visit 5). The impact on IgG levels to Spike and neutralization breadth and potency was measured. The third vaccine dose (visit 5) increased Spike IgG binding **(Figure 2)** and the plasma neutralizing activity (**Figure 5A-D**) compared to visit 4 in both groups. Whereas after the second dose there had been a clear distinction in neutralization breadth between the short and extended booster groups, following administration of a third vaccine dose the neutralizing titres against WT, alpha, delta, beta and omicron/BA.1 were very similar (**Figure 5A-D**). Importantly, a 3^rd^ vaccination with ancestral SARS-CoV-2 spike increased neutralization breadth against the most highly diverse beta and omicron/BA.1 variants (**Figure 5C**). Overall, the reduced neutralization breadth observed in the short booster group after two vaccine doses is rescued following a third vaccine dose.

### Broader neutralization is associated with have higher avidity against Spike

Antibody binding avidity has been associated with antibody maturation [21-25]. To determine whether enhanced neutralization breadth was related to higher avidity of antibody binding in the extended booster group we next measured an avidity index for samples collected 3-weeks and 6 months after the second vaccine dose (visit 3 and 4) and 3-weeks after the third vaccine dose (visit 5). The avidity was measured by comparing the area under the curve in ELISA, with and without an 8M Urea washing step. For SARS-CoV-2 naïve individuals at visit 3, the extended booster group had a significantly higher avidity index than the short booster group (**Figure 5E**). However, this difference was reduced in when comparing previously infected individuals. The avidity index 6 months post 2^nd^ vaccine (visit 4) was largely unchanged (**Figure 5F**). Following the 3^rd^ vaccination (visit 5) where neutralization breadth was similar across the two groups, the avidity index increased in both groups and the difference in avidity index between the short and extended groups was no longer significant (**Figure 5G**). The avidity index 3-weeks after 2^nd^ vaccine dose (visit 3) was correlated with the ID_50_ against each VOC (**Figure 5I**). No significant correlation was observed for WT. However, the correlation coefficient was greatest for beta and omicron/BA.1 which are most antigenically distant to the vaccine strain and suggests that avidity may be a good indicator VOC cross-reactivity. The avidity index also correlated with the % of Spike-reactive memory B cells (**Figure 5H**).

### Neutralization of BA.2 and BA.4/5 sub-lineages

Since omicron/BA.1 became the dominant global variant worldwide, several related SARS-CoV-2 omicron sub-lineages have been reported that encode unique amino acid changes in Spike. BA.2 has driven a recent wave in the UK and encodes 21 identical mutations to BA.1 (including K417N, N440K and E484A) and 8 additional mutations across NTD and RBD. BA.4 and BA.5, which encode identical Spikes proteins, are thought to be driving a 5 ^th^ wave in South Africa [26]. The BA.4/BA.5 Spike is similar to BA.2 but has additional mutations delta69-70, L452R and F486V and lacks the Q493R mutation. Emergence of these highly transmissible sub-lineages has led to an increased rate of breakthrough infections (BTIs) and, within the cohort described here, fourteen participants experienced BTI between mid-December 2021-March 2022. In mid-December BA.1 made up >80% of UK infections and the BA.1 and BA.2 sub-lineages made up 95% of infections from mid-January [27]. Therefore, BTIs in participants were presumed to be caused by BA.1 or BA.2 VOCs (visit 7). Blood was also collected from six participants 6 months post 3^rd^ dose (visit 6).

To assess susceptibility to the recently reported BA.4/BA.5 variants and to determine the impact of BA.1/BA.2 BTI on the antibody response, we measured neutralization against D614G, BA.1, BA.2 and BA.4/BA.5 following three BNT162b2 doses (at 3-weeks (visit 5) and 6-months (visit 6) post boost) and in those who subsequently experienced a BTI (3-weeks post infection (visit 7)) (**Figure 6**). Due to the smaller number of samples available, all plasma were considered together for analysis. For plasma collected 3-weeks post 3^rd^ vaccine dose (visit 5), good neutralization against BA.1, BA.2 and BA.4/5 was detected but GMTs were slightly decreased compared to D614G, with the greatest reduction being 2.2-fold against BA.4/5 (**Figure 6A**). Six months post 3^rd^ vaccine dose (visit 6), neutralization titres had decreased against all variants (**Figure 6B** and **6D**) as seen six months post 2^nd^ vaccine dose (**Figure 5**). However, in those who experienced a BA.1/BA.2 BTI (visit 7) neutralization titres increased against all variants compared to visit 5 and robust cross-neutralization of all omicron sub-lineages was detected (**Figure 6C** and **6E**). The largest fold increase in GMT following BTI was observed against BA.1 and BA.2 (**Figure 6E**).

**Figure 6:**
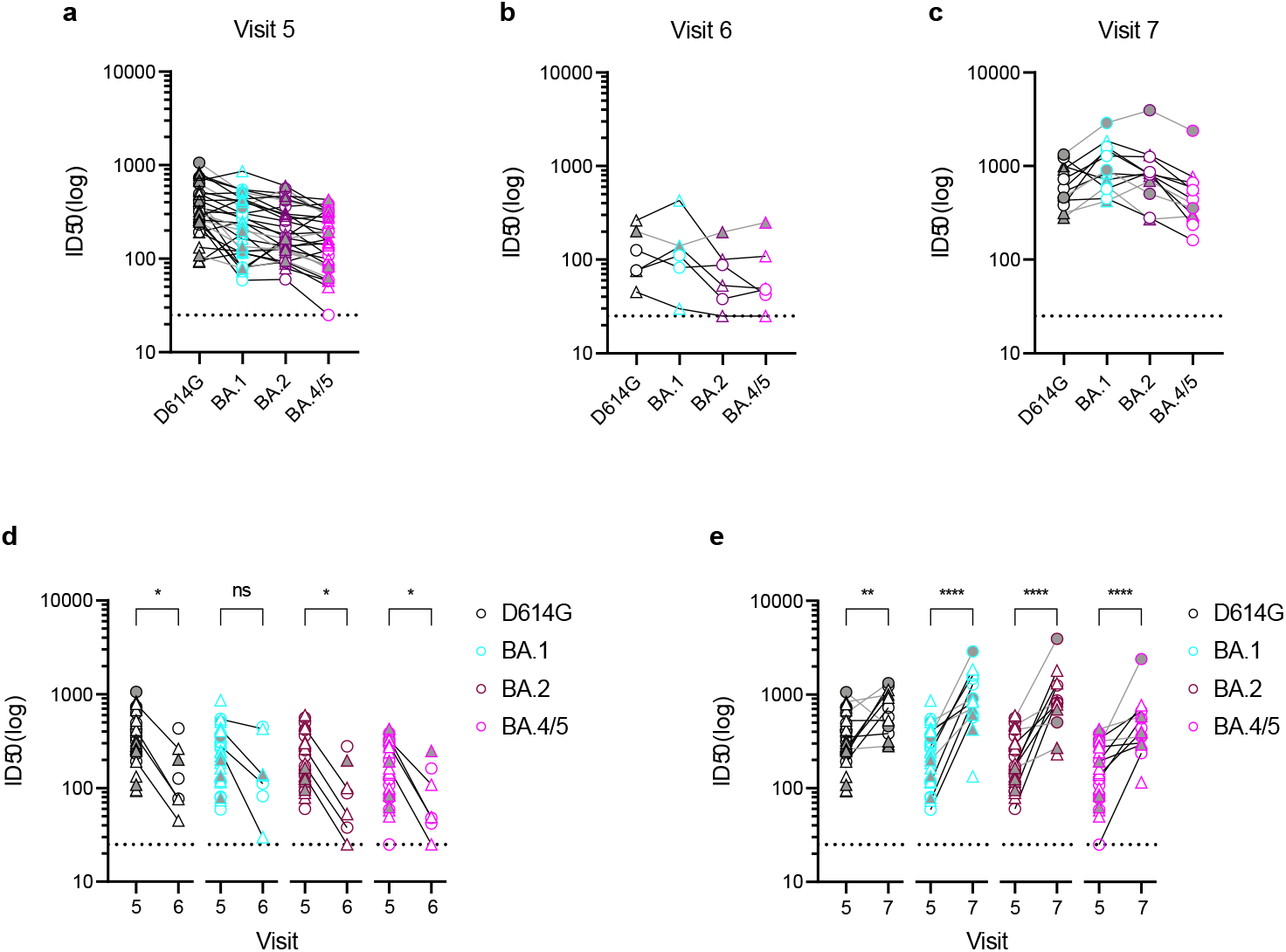
Neutralization against omicron sub-lineages following 3 vaccine doses and/or BA.1/BA.2 breakthrough infection. Neutralization breadth and potency was measured against D614G, BA.1, BA.2 and BA.4/BA.5. Analysis of sera collected **A)** 3-weeks post 3^rd^ BNT162b2 vaccine dose (visit 5), **B)** 6-months post 3^rd^ vaccine (visit 6) and **C)** following breakthrough infection (visit 7) (presumed to be BA.1 or BA.2). **D)** Comparison of titres 3-weeks (visit 5) and 6-months post 3^rd^ boost (visit 6). **E)** Comparison of titres 3-weeks post 3^rd^ boost (visit 5) and following BTI (visit 7). Samples from a single individual are joined. Short and long booster groups shown with circle and triangle symbols, respectively. Individuals infected with SARS-CoV-2 prior to vaccination are shaded grey. Based on this result, multiple Mann-Whitney tests or unpaired t tests using a two-stage linear step-up procedure of Benjamini, Krieger and Yekutieli were employed to determine significance between groups. ns P > 0.05, * P ≤ 0.05, ** P ≤ 0.01, *** P ≤ 0.001 and **** P ≤ 0.0001.

Overall, a third vaccine dose generates neutralization against omicron sub-lineages BA.1, BA.2 and BA.4/5 which is further enhanced by BA.1/BA.2 BTI.

## Discussion

Elicitation of nAbs with broad activity against both known and newly emerging variants is highly desirable and vital in controlling the current COVID-19 pandemic. Here we studied the impact of vaccine interval on the breadth and potency of the SARS-CoV-2 neutralizing antibody response. We showed that an extended interval between the 1^st^ and 2^nd^ dose of the BNT162b2 vaccine enhances neutralization breadth initially. However, a third vaccine dose generated a robust and broad neutralizing response against VOCs independent of the interval between 1^st^ and 2^nd^ vaccine doses. It has been extensively reported that ‘hybrid’ immunity, immunity derived from SARS-CoV-2 infection followed by vaccination, leads to superior neutralization following 1-dose of COVID-19 vaccine [12-16]. Our results showing higher antibody binding and neutralization in previously infected individuals receiving one BNT162b2 dose are consistent with these reports.

Here, we show than an extended interval between the 1^st^ and 2^nd^ vaccine dose enhances the immune response in four ways. Firstly, and similar to previous reports, there is an increase in IgG binding (ED_50_) to Spike in the extended booster group following the 2^nd^ immunization [23, 28, 29] and secondly, the Spike-reactive IgG bind with higher avidity [23]. Thirdly, broader neutralizing activity against VOCs (including BA.1) is generated [30, 31]. Finally, a higher percentage of Spike-reactive IgG^+^ memory B cells is detected in SARS-CoV-2 naïve individuals receiving an extended booster. Binding avidity correlated most strongly with the ID_50_ against the most antigenically distinct VOCs. The higher binding avidity in the extended group suggests that antibodies have undergone higher levels of antibody maturation [21-24]. Indeed, wider SARS-CoV-2 neutralization breadth has been associated with increased somatic hypermutation [12, 32-34]. We have recently reported that mAbs isolated from an individual receiving 2-doses of the ChadOx1 AZD1222 vaccine at a 12-week interval were more highly mutated than those isolated from SARS-CoV-2 convalescent donors and displayed greater neutralization breadth [32]. Several studies have shown that mRNA vaccination leads to a robust memory B cell response and induces persistent germinal centre reactions that continue for months after primary vaccination [12,19, 35, 36]. Therefore, a longer interval between the 1^st^ and 2^nd^ vaccine dose would allow extended germinal centre reactions to take place, leading to increased levels of somatic hypermutation and higher affinity to vaccine antigen.

Neutralization titres decreased 6 months post 2^nd^ vaccine dose [20]. However, consistent with previous research, we show that titres are boosted following the 3^rd^ BNT162b2 vaccination. Importantly, neutralization breadth increased against beta and omicron sub-lineages independent of the interval between the 1^st^ and 2^nd^ vaccine doses leading to overall similar levels of cross-neutralizing activity in both the short and extended booster groups. This increase in neutralization breadth was accompanied by an increase in the Spike-reactive IgG binding avidity in the short booster group, indicative of continued affinity maturation. Nussenzweig and co-workers identified antibody lineages that had undergone further somatic hypermutation following a 3^rd^ dose of COVID-19 mRNA vaccine and these mAbs had superior neutralization breadth and potency [37]. Relevant to the COVID-19 pandemic at the time of writing (June 2022), neutralization against the antigenically distant VOCs BA.1 [3-5, 8], BA.2 [2, 38, 39] and BA.4/BA.5 [40-42] also increased following a 3^rd^ vaccine dose in both groups.

Following BTI during the UK BA.1 or BA.2 wave, neutralization titres against D614G, BA.1, BA.2 and BA.4/BA.5 increased in this cohort independent of previous SARS-CoV-2 infection. The fold increases in GMT were larger against Omicron sub-lineages compared to against D614G. Increases in omicron sub-lineage neutralization following omicron BTI has been observed in other cohorts [43-45]. Furthermore, boosting mRNA vaccinated mice with an omicron/BA.1 based mRNA vaccine increased serum neutralization activity and protection against omicron/BA.1 [46]. Recent studies indicate that the increase in cross-neutralizing activity is mostly driven by re-activation of B cell clones initially generated through vaccination with Wuhan-1 Spike that cross-react with the VOC, with little evidence of the generation of a *de novo* VOC-specific antibody response [44, 47, 48]. These observations are contradictory to recent reports suggesting that antibody responses to omicron/BA.1 were limited following an omicron BTI in triple vaccinated donors who had been infected prior to vaccination [49].

The findings of this study are applicable in populations where COVID-19 vaccine coverage remains low and where there is limited vaccine supply. In vaccine naïve populations, an extended interval will likely be beneficial through generation of broader antibody-based immunity against VOCs in the first six months following the 2^nd^ vaccine dose. Indeed, vaccine effectiveness was shown to be significantly higher in those receiving a longer interval between mRNA doses [50, 51]. However, whilst there is broader activity following the 2^nd^ vaccine dose in the extended group, there would be a longer period following the 1^st^ vaccine in which antibody titres will remain low and narrow which in turn may lead to a higher risk of infection during this period. Importantly, despite the interval between the 1^st^ and 2^nd^ vaccine doses, a third vaccine dose increases neutralization against the current VOCs including omicron sub-lineages BA.1, BA.2, BA.4 and BA.5.

In summary, in a SARS-CoV-2 naïve population, an extended interval between the 1^st^ and 2^nd^ doses of BNT162b2 leads to an increased neutralization breadth, Spike binding avidity and frequency of Spike-reactive memory B cells. However, this advantage is lost when a 3^rd^ vaccine dose is administered 6-8 months later where broad neutralization against current omicron sub-lineages is observed in both groups. These findings provide insights into optimizing vaccine regimens to provide the greatest neutralization breadth.

## Limitations of the study

A limitation of this study is the relatively small sample size in each group which limits the power of the statistical analysis. The impact on cell-mediated immune responses has not been investigated. However, Hall *et al* have previously show that no significant differences in Spike-specific polyfunctional CD4+ T cell responses between the short and extended groups [52] whereas Payne et al reported higher levels of these cells in the extended group [28]. This study does not assess whether the broader neutralization response in the extended-group reduced breakthrough infections.

## Materials and Methods

### Ethics

This study used human samples collected with written consent as part of a study entitled “Antibody responses following COVID-19 vaccination”. Ethical approval was obtained from the King’s College London Infectious Diseases Biobank (IBD) (KDJF-110121) under the terms of the IDB’s ethics permission (REC reference: 19/SC/0232) granted by the South Central – Hampshire B Research Ethics Committee in 2019.

### Protein expression and purification

Recombinant Spike and nucleoprotein for ELISA were expressed and purified as previously described [17, 53].

### ELISA (Spike and nucleoprotein)

96-well plates (Corning, 3690) were coated with antigen at 3 μg/mL overnight at 4°C. The plates were washed (5 times with PBS/0.05% Tween-20, PBS-T), blocked with blocking buffer (5% skimmed milk in PBS-T) for 1 h at room temperature. Serial dilutions of plasma in blocking buffer were added and incubated for 2 hr at room temperature. Plates were washed (5 times with PBS-T) and secondary antibody was added and incubated for 1 hr at room temperature. IgG was detected using Goat-anti-human-Fc-AP (alkaline phosphatase) (1:1,000) (Jackson: 109-055-098). Plates were washed (5 times with PBS-T) and developed with AP substrate (Sigma) and read at 405 nm.

### Avidity ELISA

The ELISA was carried out as described above. A 4-point titration (starting at 1:25, 1:4 dilution series) was used. After incubation of plasma, one half of the plate was incubated with 8M Urea and the other half incubated with PBS for 10mins before washing 5-time with PBS-T. The area under the curve was determined in Prism (Log dilution). The avidity index was calculated using the following formula:

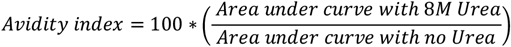

### Neutralisation assay with SARS-CoV-2 pseudotyped virus

Pseudotyped HIV-1 virus incorporating the SARS-CoV-2 Spike protein (either wuhan-1, D614G, alpha (B.1.1.7), beta (B.1.351), delta (B.1.671.2), mu (B.1.621), omicron (BA.1, BA.2 or BA.4/5) were prepared as previously described [17, 54]. Viral particles were produced in a 10 cm dish seeded the day prior with 5×10^6^ HEK293T/17 cells in 10 ml of complete Dulbecco’s Modified Eagle’s Medium (DMEM-C, 10% FBS and 1% Pen/Strep) containing 10% (vol/vol) foetal bovine serum (FBS), 100 IU/ml penicillin and 100 μg/ml streptomycin. Cells were transfected using 90 μg of PEI-Max (1 mg/mL, Polysciences) with: 15μg of HIV-luciferase plasmid, 10 μg of HIV 8.91 gag/pol plasmid and 5 μg of SARS-CoV-2 spike protein plasmid [55, 56]. The supernatant was harvested 72 hours post-transfection. Pseudotyped virus particles was filtered through a 0.45μm filter, and stored at -80°C until required.

Serial dilutions of plasma samples (heat inactivated at 56°C for 30mins) were prepared with DMEM media (25µL) (10% FBS and 1% Pen/Strep) and incubated with pseudotyped virus (25µL) for 1-hour at 37°C in half-area 96-well plates. Next, Hela cells stably expressing the ACE2 receptor were added (10,000 cells/25µL per well) and the plates were left for 72 hours. Infection levels were assessed in lysed cells with the Bright-Glo luciferase kit (Promega), using a Victor™ X3 multilabel reader (Perkin Elmer). Each serum sample was run in duplicate and was measured against the five SARS-CoV-2 variants within the same experiment using the same dilution series.

### FACS analysis of Spike-specific memory B cells

Flow cytometry of cryopreserved PBMCs was performed on a BD FACS Melody as previously described. SARS-CoV-2 Wuhan-1 Spike was pre-complexed with the streptavidin fluorophore (Alexa-488) at a 4:1 molar ratio prior to addition to cells. PBMCs were incubated with Fc block for 15 minutes at 4°C. PBMCs were stained with anti-CD3-BV510 (Biolegend), anti-CD19-PerCP-Cy5.5 (Biolegend), anti-IgD-Pacific Blue (Biolegend), anti-CD27-BV785 (Biolegend), anti-CD38-APC-Cy7 (Biolegend), anti-CD20-APC (Biolegend), anti-IgG-PE-Cy7 (BD Biosciences), anti-IgM-PE (Biolegend) and Spike-Alexa Fluor 488 for 1 hour at 4°C. PBMCs were washed with PBS and stained with live/dead for 1 hour at 4°C.

## Data Availability

All data produced in the present work are contained in the manuscript

## Acknowledgements

We thank Philip Brouwer, Marit van Gils and Rogier Sanders for the Spike protein construct, Leo James and Jakub Luptak for the N protein, Wendy Barclay for providing the B.1.617.2, BA.1, BA.2 and BA.4/BA.5 Spike plasmids and James Voss and Deli Huang for providing the Hela-ACE2 cells.

