## Supplemental information for "The effect of Omicron breakthrough infection and extended BNT162b2 booster dosing on neutralization breadth against SARS-CoV-2 variants of concern"

### Extended BNT162b2 booster dosing generates a broader neutralizing antibody response against SARS-CoV-2 variants of concern

Carl Graham[1*], Thomas Lechmere[1*], Aisha Rehman[1], Jeffrey Seow[1], Ashwini Kurshan[1], Isabella Huettner[1], Thomas J.A. Maguire[1], Jerry Tam[1], Daniel Cox[1], Christopher Ward[1], Mariusz Racz[2], Anele Waters[2], Christine Mant[3], Michael H. Malim[1], Julie Fox[1], Katie J. Doores[1]#

[1] Department of Infectious Diseases, School of Immunology & Microbial Sciences, King's College London, London, UK.

[2] Harrison Wing, Guys and St Thomas' NHS Trust, London, UK.

[3] Infectious Diseases Biobank, Department of Infectious Diseases, School of Immunology and Microbial Sciences, King's College London, London, UK.

* These authors contributed equally

**Supplementary Figure S1: SARS-CoV-2 Spike-reactive memory B cells measured before and after vaccination. A)** Example FACS gating for pre-vaccination sample (visit 1) and from post-vaccination sample (visit 3) from SARS-CoV-2 naïve individual. **B)** Frequency of memory B cells in the total B cell population at visit 1 and visit 3 for matched donors.

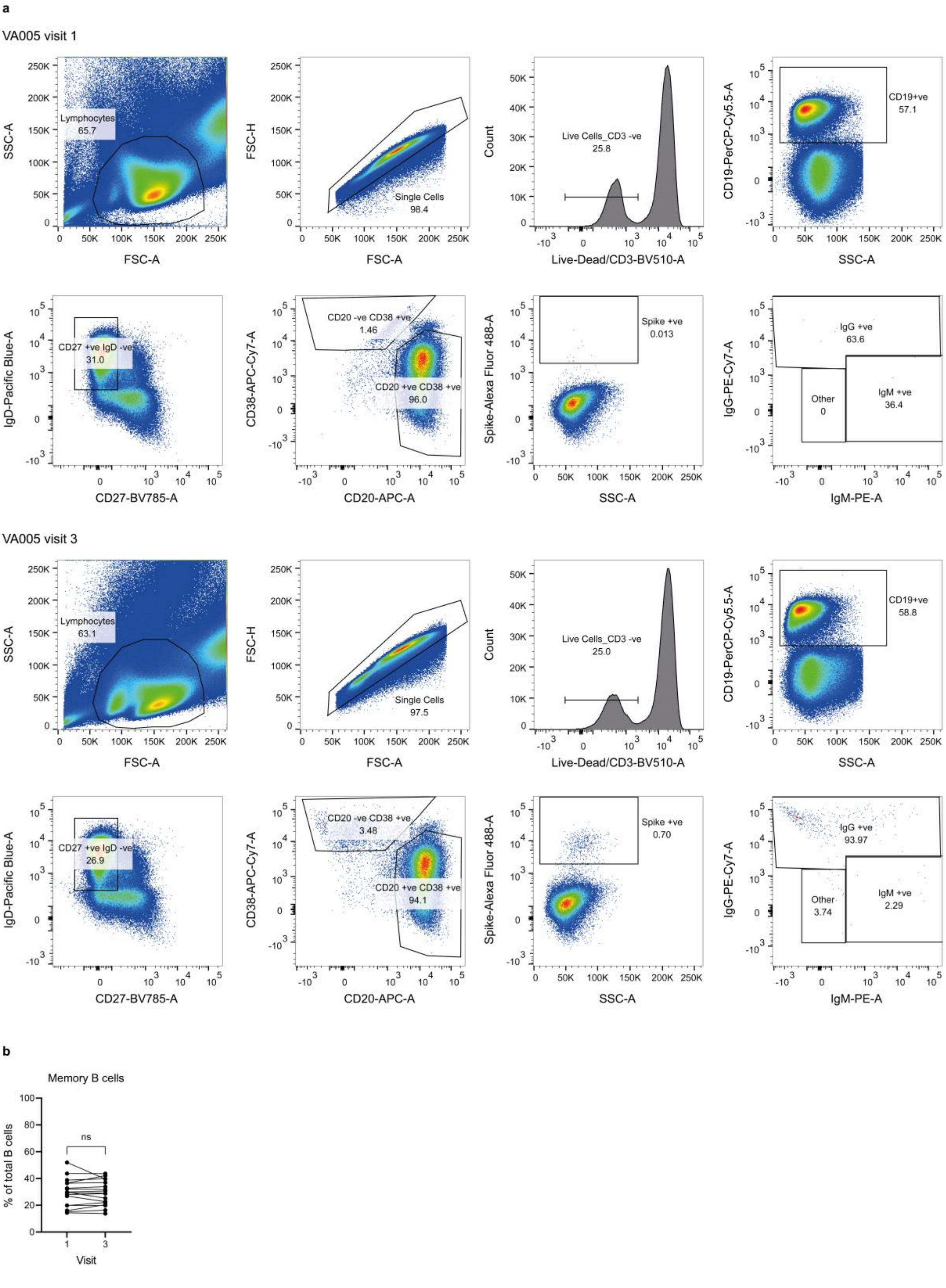
a
VA005 visit 1
Lymphocytes
65.7
SSC-A
FSC-A
FSC-H
Single Cells
98.4
Count
Live Cells_CD3 -ve
25.8
Live-Dead/CD3-BV510-A
CD19-PerCP-Cy5.5-A
CD19+ve
57.1
SSC-A
IgD-Pacific Blue-A
CD27 +ve IgD -ve
31.0
CD27-BV785-A
CD38-APC-Cy7-A
CD20 -ve CD38 +ve
1.46
CD20 +ve CD38 +ve
96.0
CD20-APC-A
Spike-Alexa Fluor 488-A
Spike +ve
0.013
IgG-PE-Cy7-A
IgG +ve
63.6
Other
0
IgM +ve
36.4
IgM-PE-A
VA005 visit 3
Lymphocytes
63.1
Single Cells
97.5
Live Cells_CD3 -ve
25.0
CD19+ve
58.8
CD27 +ve IgD -ve
26.9
CD20 -ve CD38 +ve
3.48
CD20 +ve CD38 +ve
94.1
Spike +ve
0.70
IgG +ve
93.97
Other
3.74
IgM +ve
2.29
b
Memory B cells
% of total B cells
ns
Visit
